# The Care Burden and Influencing Factors of Family Caregivers of Older Adults with Dementia

**DOI:** 10.1101/2023.02.24.23286414

**Authors:** Laiyou Li, Ning Sun, Hongyin Wang, Chaoyan Fan, Hongyu Li, Shuang Yang

## Abstract

**Objective:** To analyze the current state of care burden of dementia caregivers and determine the influencing factors.

**Methods:** Using the questionnaire survey method, 600 family caregivers of older adults with dementia in five communities in Ningbo (China) were investigated from March to August 2022.

**Results:** The average score of care burden of dementia was 52.36 ± 12.08. The total score of social support for older adults with dementia ranged from 19 to 56, with the average score being 34.79 ± 9.54. The total score of social support and the score of each dimension were negatively correlated with the total score of the care burden and the score of each dimension, with the correlation coefficient being between 0.490 and 0.689 (p <0.05). Six factors affecting the burden of care for family caregivers of older adults with dementia were total social support score, dementia level, monthly family income, years of care, educational level, and the duration of daily care for older adults with dementia (all p <0.001).

**Conclusion:** The care burden of older adults with dementia is significant, with many influencing factors. Therefore, it is necessary to relieve the care burden of family caregivers through education support programs, self-efficacy training of caregivers, breathing care services, and psychological counseling to improve the quality of life, and relieve the pressure of social counseling and social support.

## 1. Introduction

Dementia refers to a common group of advanced brain dysfunctions caused by chronic or progressive organic structural damage in old age. It is mainly manifested as persistent, comprehensive intelligence decline, behavioral abnormalities, and a loss of ability to live and work independently (1). At present, dementia has become the fourth cause of death in developed countries after cardiovascular disease, malignant tumors, and stroke (2). By the end of 2022, there were about 13 million older adults with dementiain China, accounting for about 25 percent of the world’s total population with this condition (3)It is predicted that in 2040, the number of older adults with dementia in China will reach about 22 million, which is the sum of older adults with dementia in all developed countries. Therefore, the care of older adults with dementia has become a prominent social problem in the aging population in China.

The families of dementia patients have a heavy burden, spending 26,000 to 35,000 yuan per person per year (5). The quality of care plays a pivotal role in the quality of life of older adults with dementia, and the physical and mental health of the caregivers is an important guarantee of the quality of care. Older adults with dementia can not fully care for themselves in daily life and require long-term care due to the deterioration of the condition and the accompanying mental and behavioral abnormalities. Long-term care not only makes the caregivers physically tired but also damages their mental health, further increasing the burden of care. The harm of caregiver burden should not be underestimated. Long-term lack of effective response will lead to negative consequences from overwork, causing chronic fatigue syndrome in mild cases, and sudden death in severe cases. Many people are likely to become a caregiver at some point in their lives, and caring about the physical and mental health of the caregiver is imperative to the future. to pay attention to our tomorrow. This study explores the burden of care of family caregivers with dementia, and the factors influencing this burden in order to encourage caregivers to adopt active responses, actively accept and make full use of social support, relieve fatigue, ensure physical and mental health, and ensure the quality of care.

Foreign research on care burden began in the 1970s and entered the stage of rapid development in the early 21st century (6). In 2013, the World Health Organization-World Mental Health Alliance survey of 19 participating countries demonstrated that 40% of the countries’ caregivers had a different degree of care burden for the caregivers due to care time and economic factors; in terms of subjective burden, the psychological burden due to the disability of older adults was also noted (7). In foreign countries, Zarit and others first started studying the caregivers of Alzheimer’s disease patients, and then the researchers gradually conducted studies with different populations, mainly focusing on the caregivers of older adults with Alzheimer’s disease patients (8-10).

Studies have shown that caregiver burden includes physical, psychological/emotional social and economic components (11,12). Different types of care burden have different factors and are related to a caregiver’s environmental and social factors. At present, it is generally believed that the factors influencing caregiver burden include: daily living ability and mental behavior symptoms of older adults, physical strength required for caring for older adults with dementia, caregivers’ responses to dementia behavior, caregivers’ lack of social activities, and economic stress (13,14). It is estimated that 20% to 50% of family caregivers are depressed due to the heavy burden of care, which is 2 to 3 times (14,15) that of the general population (14,15). Studies have shown that care burden is positively associated with factors such as lack of educational support, care skills and coping strategies, time spent to maintain activities of daily living in older adults with dementia, and lack of general support (11,16,17). Caregiver burden is also related to the recipient, including the severity of dementia and dementia-related psychobehavioral symptoms (18,19). Educational support programs, caregiver self-efficacy training, wheezing care services, psychological counseling, social support, and the establishment of a dementia social network for dementia caregivers can all alleviate the caregiver burden (20,21).

In the 1990s, China began to carry out relevant studies on the care burden, which started relatively late in other foreign countries. However, domestic studies on care burden mostly focus on cross-sectional studies of small and specific groups, mainly focusing on caregivers of older patients with Alzheimer’s disease (22,23). Some scholars studied 100 family caregivers of older adults with dementia and concluded that the top three care burdens were emotional burden, social activities, and the burden of time dependence (24). At the same time, Zhu Rong (25) and other scholars investigated and analyzed 122 caregivers who care for older adults with dementia and found that the overall care ability of family caregivers is relatively low, and their educational level, physical health, awareness of the disease and coping methods are all factors affecting the care ability. Zhang Ji’an study (26) of eight family caregivers found that family caregivers had no time and energy to take care of their children or to engage in normal social interaction to promote individual growth, in the process of care, need to endure the family of forgotten members and care work. Zhuang Shuting (27) adopted a case study method integrating literature and in-depth interviews to study coping strategies from the perspective of family caregivers, and put forward feasible suggestions to help them reduce their care burden. Tang Yong (28) proposed that the care of older adults is mainly provided by family members, and that care for older adults takes most of a caregiver’s normal social interaction time. Less communication between family members was also observed, along with decreased caregiver time for children. Further, caregivers are sometimes unable to continue working, affecting family income and quality of life. The older population continues to rise, and require more detailed and comprehensive care. Therefore, a comprehensive understanding of the current situation of family caregivers’ care burden and its influencing factors may allow for effective intervention measures that prevent or reduce caregiver fatigue, prevent physical and mental health conditions, ensure the quality of care for older adults with dementia, and improve the quality of life of older adults with dementia.

## 2. Methods

### 2.1 Study design

A correlational, cross-sectional design was adopted, and questionnaires were used for data collection.

### 2.2 Setting and sample

In this study, the convenience sampling method used the list of older adults people with dementia provided by clinicians in five communities of Ningbo from April to July 2022. A total of 600 older adults with dementia were identified to conduct questionnaires on their home caregivers. Participant inclusion criteria were as follows: 1) Family caregivers of older adults with dementia who were diagnosed by clinicians; 2) being in a caregiving position for at least 1 year; 3) living in a community with older adults with dementia; 4) be over 18 years old, conscious, and able to voluntarily participate in the study. The exclusion criteria included: 1) The caregiver has a cognitive impairment; 2) has severe heart, liver, lung, and brain dysfunctions.

### 2.3 Sample size calculation

According to the sample content should be 10∼20 times of the study variable, the number of items in the care burden scale is 24. Considering the insufficient sample size caused by questionnaire misfilling, missing filling, and invalid questionnaires, the sample size was increased by 20%. The sample size of this study is calculated as follows: N= [24 (10∼20)] (1 + 20%), and the required sample size was 288-576 people.

### 2.4 Data collection

Two investigators underwent unified training for data collecting. Data was collected through face-to-face interviews, after participants engaged in a scheduled meeting arranged by the managers in the community. The investigators explained the research objectives and methods to individuals who met the inclusion criteria and obtained informed consent and cooperation from participants. Consenting participants received an envelope containing a packet with the questionnaires. Participants completed the questionnaires immediately upon receipt and placed them in the envelope for collection by the investigators. To ensure anonymity, each completed questionnaire was assigned a code number.

### 2.5 Instruments

Four structured questionnaires were used for the data collection. The general information questionnaire for family caregivers of older adults with dementia consisted of general information, including gender, age, occupation, marital status, educational level, monthly income, medical insurance payment method, number of joint caregivers, relationship with older adults with dementia, years of care, length of care, etc. The Mini-Mental-State-Examination (MMSE), designed in 1975 by Folstein et al., is a clinical tool for screening Alzheimer’s disease patients. It is a widely used cognitive function screening scale at home and abroad, and it is currently the most influential screening tool for dementia (29). There were 19 items with a total score of 30, and the subject’s MMSE score of 24. Considering the patient had dementia or other diseases with a cognitive function component, but the subject score was closely related to their education level, and the cut-off value of dementia had different educational levels. It has been reported in the literature that the standard cut-off value was set as 24 points, 17 points for the illiterate (uneducation) group, 20 points for the primary school (6 years of education) group, and 24 points for the middle school or above group. The Cronbach’s α was 0.83 (30).

The caregiver burden questionnaire (Caregiver Burden Inventory, CBI), translated by Taiwan scholar Chou et al, (31), was used to investigate the home caregiver care load. It contains 5 dimensions (time-dependent, development-limited, physical, social, and emotional load) and 24 items. The scoring method was 0 to 4, and the scoring range was 0 to 96. Higher scores indicate a heavier caregiving load. The questionnaire CVI was 0.95 and Cronbach’s α 0.85. The Cronbach’s α of this questionnaire was 0.90.

The Social Support Scale (SSRS), developed by the scholar Xiao Shuiyuan (32), is used to measure the level of social support obtained by a certain group of people. The scale’s Cronbach’s α coefficient is 0.94, the reliability is high (33), and the domestic norm is 44.38 ± 8.38 with a score of 34. The scale consists of 10 entries, which can be divided into 3 dimensions. The objective support dimension consists of items 2,6, and 7, the subjective support dimension consists of items 1,3,4, and 5, respectively, and the utilization dimension of support consists of items 8,9, and 10. The total score is 12 to 66, and a lower score indicates a lower level of social support. Regarding social support level, 22 indicates a low level, 23 to 44 indicates a middle level, and 45 to 66 indicates a high level.

### 2.6 Ethical considerations

This study and the gathering of data were approved by the XX University Institutional Review Board. All participants provided written informed consent before participating in this study. Two researchers were responsible for informing participants, both in writing and orally, about the purpose of the study and the data gathering. The participants were informed that the survey was completely voluntary and that withdrawal from the study was possible at any time, without any negative consequences.

### 2.7 Data Analysis

Questionnaires were collated, checked, and entered by two people, and the SPSS26 was used for statistical analysis (results were considered significant if p <0.05). The specific methods were as follows:

1. Statistical description: in the general data, care burden and social support data of older adults with dementia and their caregivers are given; the counting data are described by frequency and percentage; the measurement data of normal distribution are described by the mean ± standard deviation, and the measurement data of biased distribution are described by the median and interquartile spacing (M, P75-P25; x ±s).
2. correlation analysis: Pearson correlation was used to analyze the correlation between social support and care burden scores for caregivers of older adults with dementia.
3. Multiple linear regression analysis: Multiple linear regression analysis with the dependent variable, statistically significant variables and the total score of social support in the univariate analysis as independent variables, and multiple linear gradual regression analysis was conducted to explore the influencing factors of care burden for for caregivers of older adults with dementia.

## 3. Results

### 3.1 Participant demographics

A total of 600 questionnaires were distributed and 520 valid questionnaires were recovered, with an effective recovery rate of 86.7%. Of the 520 caregivers in this study, 85 were male (16.3%), and 435 (83.7%) were female; the age range was between 24 and 87 years (mean = 58.38 ± 12.18), with most being between 40 and 59 years; 418 (80.4%) had completed primary school, 73 were students (14.0%), 23 (4%) were in high school and technical secondary school, and 6 cases (1.2%) were in junior college and above; most were married (88.8%). Detailed data are shown in Table 1.

**Table 1.**
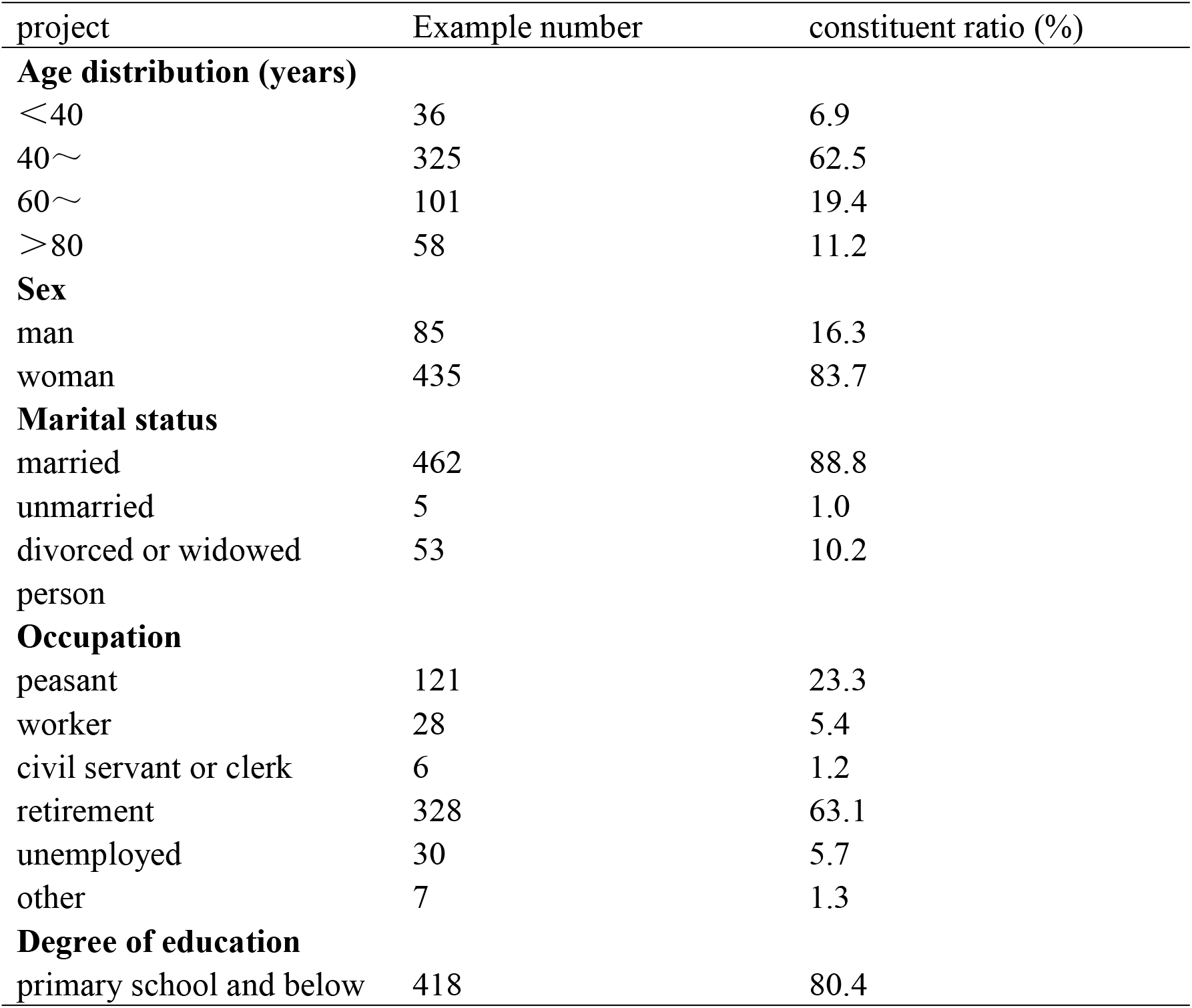
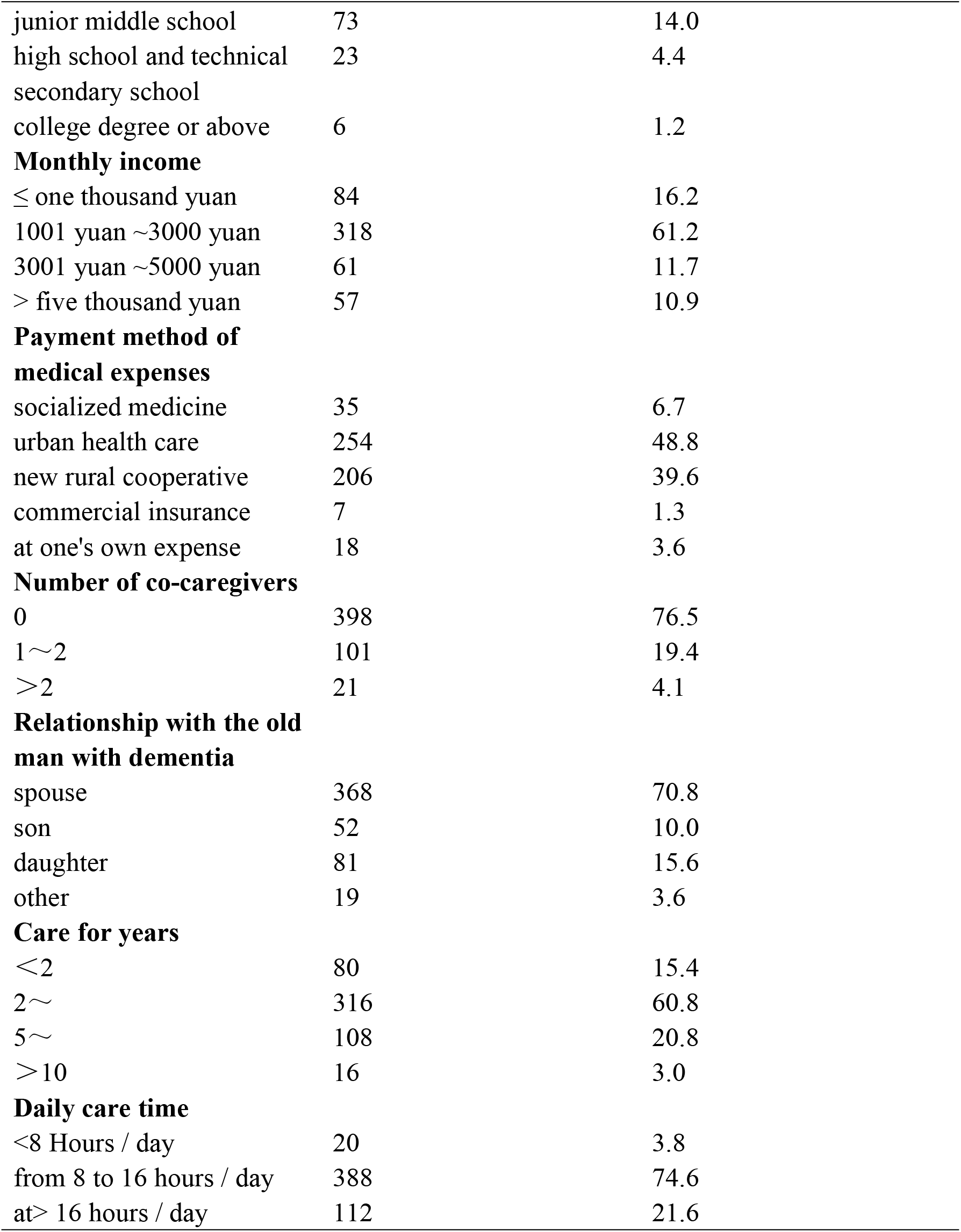
Demographic Data of family caregivers of older adults with dementia (n=520)

### 3.2 Family caregivers of older adults with dementia

The average burden of family caregivers for older adults with dementia was 52.36 ± 12.08, and the top three burdens were emotional (16.28 ± 5.21), social (14.52 ± 2.96), and time dependence (11.98 ± 3.78), as shown in Table 2.

**Table 2.** Care load scores of elderly family caregivers with dementia (n=520)

| project | Theoretical score | Theoretical Score (x ± s) | scoring average (%) |
| --- | --- | --- | --- |
| total points | 0~96 | 52.36±12.08 | 58.10 |
| time dependence | 0~20 | 11.98±3.78 | 60.25 |
| Restricted development | 0~20 | 7.46±3.52 | 36.98 |
| Body sex | 0~16 | 6.49±3.19 | 40.98 |
| koinotropic type | 0~16 | 14.52±2.96 | 79.80 |
| emotionality | 0~24 | 16.28±5.21 | 61.64 |

### 3.3 Social support for the family caregivers of older adults with dementia

The total score of social support for caregivers of older adults with dementia ranged from 19 to 56, with the average score being 34.79 ± 9.54. The objective support dimension score ranged from 3 to 18, and the average score was 9.67 ± 3.23. The subjective support dimension score ranged from 10 to 32.Between, the average score is (18.62 ± 5.12); the support utilization dimension score is between 3 and 12, with the average score being (6.50 ± 2.54), as shown in Table 3.

**Table 3.** Score of social support among family caregivers with dementia (n=520)

| Project | Number of entries | least value | crest value | Theoretical Score (x ± s) |
| --- | --- | --- | --- | --- |
| Objective support | 3 | 3 | 18 | 9.67±3.23 |
| Subjective support | 4 | 10 | 32 | 18.52±5.12 |
| Usage of support | 3 | 3 | 12 | 6.50±2.54 |
| Total social support score | 10 | 19 | 56 | 34.79±9.54 |

### 3.4 Correlation of care burden with social support

The results showed that the total score of social support and the score of each dimension were negatively correlated with the total score of caring burden and the score of each dimension, The correlation coefficient was between 0.490 and 0.689 (P <0.05), as shown in Table 4.

**Table 4.** Correlation of care burden and social support for family caregivers with dementia (n=520)

| dementia (n=520) |  |  |  |  |  |
| --- | --- | --- | --- | --- | --- |
| project | time dependence | Restricted development | Body sex | koinotropic type | emotionality |
| Objective support | -0.548** | -0.596** | -0.588** | -0.524** | -0.536** |
| Subjective support | -0.636** | -0.639** | -0.660** | -0.653** | -0.619** |
| Usage of support | -0.501** | -0.490** | -0.509** | -0.516** | -0.566** |
| social support | -0.660** | -0.676** | -0.689** | -0.628** | -0.626** |

\*\* $P < 0.01$

### 3.5 Factors influencing care burden for caregivers of older adults with dementia

Multiple linear stepwise regression analysis was used to further explore the influence of each variable on the care burden of caregivers of older adults with dementia, with caregiver care burden scored as dependent variables, statistically significant variables, and total score of social support as independent variables, and a linear stepwise regression model (α =0.05, α =0.1). The assignments of independent variables are shown in Table 5.

**Table 5.**
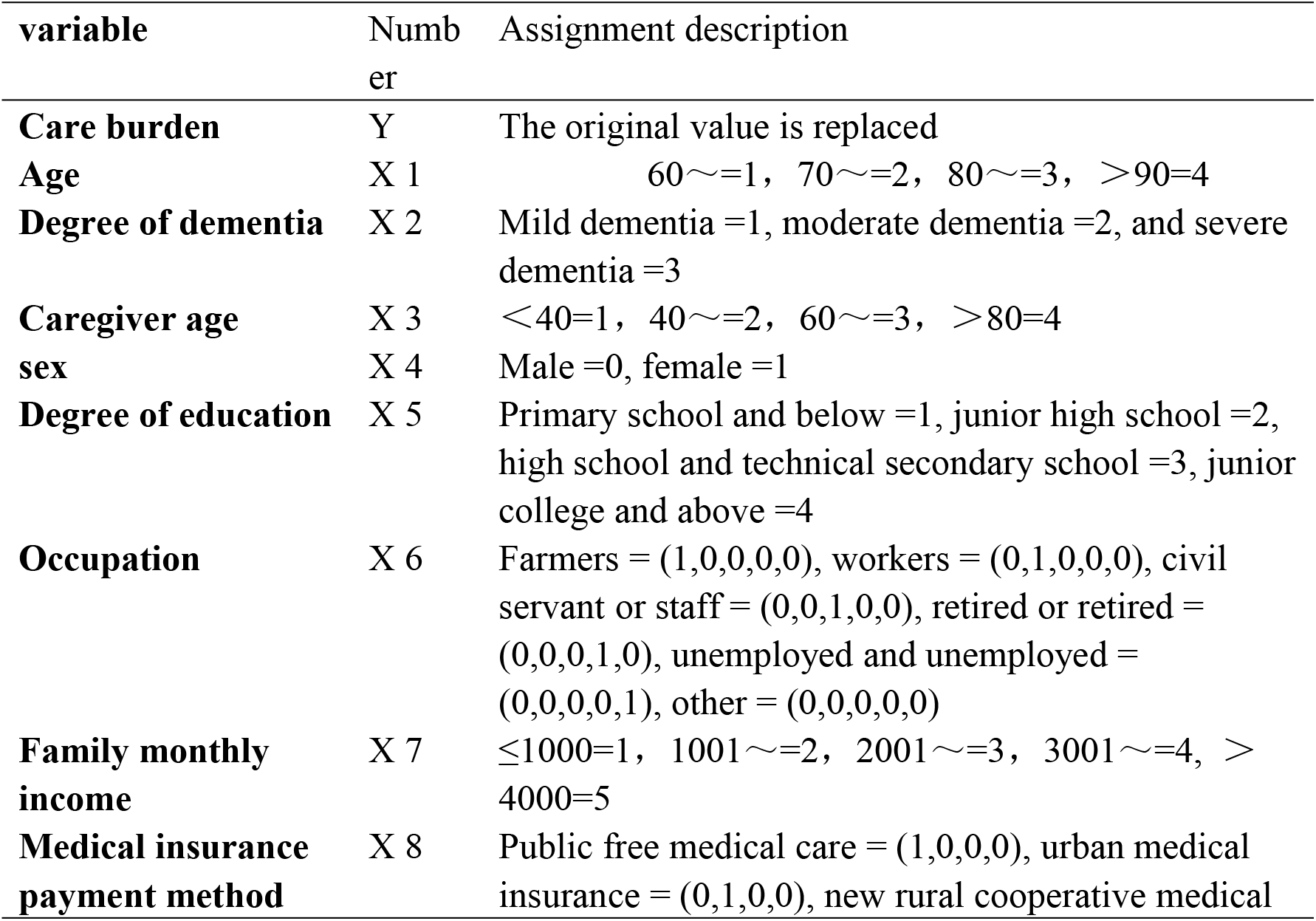
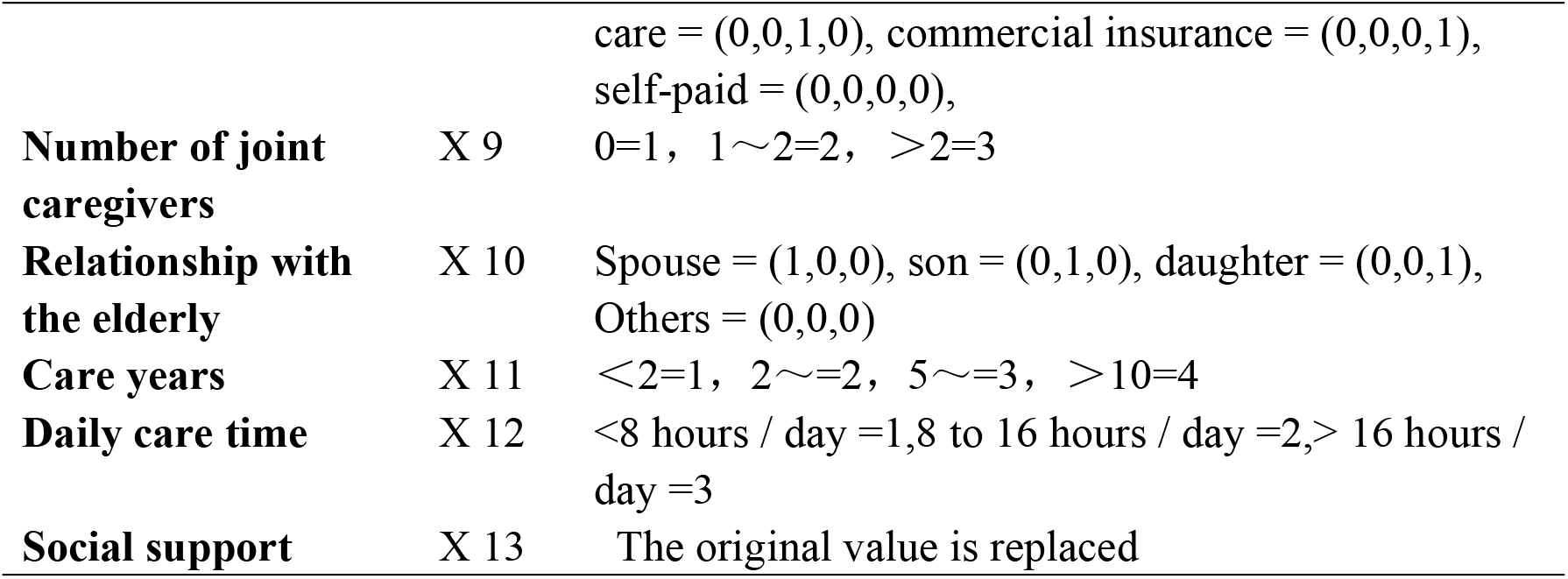
Assignment table of multiple linear regressors (n=520)

The results of multiple linear regression analysis showed that six variables were entered into the regression model: total score of social support, degree of dementia, monthly family income, years of care, education level, and time spent on daily care for older adults with dementia. The model was tested for significance using ANO variance and showed F=53.842, P <0.001, which was statistically significant. See Table 6 for details.

**Table 6.** Multiple linear regression analysis of care burden among elderly caregivers with dementia (n=520)

| **argument** | $\beta$ | S E | $\beta'$ | t | *P price* |
| --- | --- | --- | --- | --- | --- |
| **constant** | 37.589 | 6.852 |  | 5.369 | <0.001 |
| **social support** | -6.028 | 0.895 | -0.268 | -5.632 | <0.001 |
| **Degree of dementia** | 4.632 | 0.913 | 0.198 | 4.756 | <0.001 |
| **Family monthly income** | -4.536 | 0.918 | -0.269 | -4.014 | <0.001 |
| **Care for years** | 4.798 | 1.056 | 0.268 | 4.326 | <0.001 |
| **degree of education** | -4.236 | 1.269 | -0.168 | -3.115 | 0.001 |
| **Daily care time** | 2.168 | 1.326 | 0.088 | 2.089 | 0.046 |

## 4. Discussion

### 4.1 General data analysis of caregivers of older adults with dementia

Results demonstrated that caregivers of older adults with dementia were often women, at 83.7%, and they were related to the role that women play in society. Age was mainly concentrated in the 40-59 years age group, similar to the study by Wang Jing et al. Most caregivers were married, possibly because the caregivers of older adults with dementia in this survey were mainly middle-aged people. The monthly family income mostly fell between 1001∼3000 yuan, which may be related to caregivers’ limited time and ability to work. Most caregivers care for older adults alone, or with their spouses, which may be because most young adults go out to work and have no time and energy to take care of older adults.

### 4.2 Family caregivers of older adults with dementia have a heavy burden of care

In this study, the average burden of family caregivers of older adults with dementia was 52.36 ± 12.08, which was a high burden; the top three were emotional (16.28 ± 5.21), social (14.52 ± 2.96), and time dependence (11.98 ± 3.78), which were similar to results of Xiao Wenyu et al. (37). Society should be timely in providing appropriate help and support, care knowledge and care skills guidance, as well as evaluating caregivers; physical and mental health, finding existing health problems, and offering psychological counseling to reduce the overall burden of care. This can not only improve the quality of life of caregivers but can better equip caregivers to care for older adults with dementia, as is demonstrated by various domestic studies (38,39).

### 4.3 Association of care burden and social support in dementia

The results of the correlation analysis showed that social support was negatively associated with care burden, that is, caregivers with higher social support had less of a care burden, which is consistent with the findings of Lu Xueqin et al. (40). This may be because good social support has the role of relieving pressure. When family caregivers provide care services, if they can receive all aspects of objective material support or psychological comfort, they can effectively solve the caregivers’ life difficulties and relieve negative emotions.

### 4.4 Analysis of factors of dementia

#### 4.4.1 Degree of dementia

The results of multivariate analysis showed that the more severe the dementia of older adults, the heavier the care burden of the caregivers, and the difference was statistically significant (*p*<0.05). The correlation coefficient was 0.198, which is consistent with the results of previous studies (35). This may be because older adults with more severe dementia have more needs that must be met. In addition, older adults with dementia who are completely disabled depend entirely on the caregiver. This adds a greater burden on the caregiver who must solely provide physical mobility assistance to older adults.

#### 4.4.2 Family monthly income

The results of the multivariate analysis demonstrated that caregivers with lower monthly household income have a heavier care burden, with a statistically significant difference (p<0.05), and a correlation coefficient of -0.269. This is consistent with previous findings (36). This may be because most older adults with dementia have other diseases that require long-term medication, and the caregiver family needs to bear this high medical cost. In order to care for older adults, caregivers generally suspend work or quit their previous jobs, reducing the family’s finances. In addition, caregivers have little source of income, with health insurance only minimally covering medical costs.

#### 4.4.3 Care for years

The results of multivariate analysis showed that the longer the period of care, the heavier the burden of care (p <0.05), with a correlation coefficient of 0.268, consistent with the study results of domestic scholar Peng Yunhui et al. (41). This may be because the longer care time leaves the caregivers without rest time, which can partly damage the physical health of the caregivers. In addition, the caregivers have no personal free time, and no opportunity to participate in recreational activities. Caregivers are also prone to anxiety, depression, irritability, other negative psychological moods, and long time of negative psychological states that can negatively affect the caregiver’s mental health.

#### 4.4.4 Degree of education

The results of the multivariate analysis showed that the lower the education level, the heavier the care burden, and the difference was statistically significant (p <0.05), with a correlation coefficient of -0.168, which was similar to the results of the study by Ge Xiaohua et al. (39). Caregivers with higher educational levels have greater care knowledge and skills learned through the Internet, TV, mobile phones and other devices, and they also know how to search for care problems through the Internet. In addition, the caregivers can skillfully use WeChat, QQ, and other social software, and can communicate with family and friends, which can not only strengthen communication with the outside world but also relieve negative emotions.

#### 4.4.5 Daily care time

Multivariate analysis results show that the longer the daily care for older adults with dementia, the heavier burden, with the difference being statistically significant (p <0.05), and the correlation coefficient being 0.088. The main daily care of older adults, dementia need to take care of the dementia more needs, need to take longer time, caregivers have no breathing and rest time and opportunity, aggravating the burden of the body, is also more likely to accumulate negative emotions, physical and mental health may be damaged.

#### 4.4.6 Social support

The results showed that social support could negatively predict the caregiving burden of caregivers, with a correlation coefficient of -0.268, which is similar to the results of the study by Lu Xueqin et al. (40). This may be because caregivers can relieve the financial stress of care if they receive material support from family, friends or society. At the same time, if the caregiver can get relief and support from all sides, the caregiver’s psychology can be relieved. In addition, if the caregivers can get substantial help from their families and other social groups, they can relieve the burden of taking care of older adults with dementia, and can also share the care task, allowing caregivers the time and opportunity to rest.

### 4.5 Limitations and Future Research

The first limitation of the present study is that we used convenience samples and self-reports; thus, common method variance could bias our results. Therefore, we suggest a longitudinal multi-source study to test the causal effects of the variables. Second, the data we obtained were only from one city in China; therefore, the results may not be generalized to more cities. Thus, it would be interesting for researchers to involve a larger sampling range to check the proposed model in our study.

## 5.Conclusion

Family caregivers of older adults with dementia are under great pressure in various aspects. To mitigate this pressure, attention should be given to perfecting the social security system, strengthening nursing services, promoting psychological knowledge skills, increasing economic subsidies, using social workers, and enhancing social interaction to enhance nursing knowledge. This can improve the quality of care and may alleviate family caregivers’ care burden, improve their quality of life, and alleviate the pressure of social endowment.

## Data Availability

Data are available from the Ningbo college of health sciences Ethics Committee for researchers who meet the criteria for access to confidential data

## Author Contributions

The authors were responsible for the paper as follows: LYL, conception, design, analysis, and data interpretation, drafting the manuscript, revising the manuscript, and its final approval; NS, acquisition of data, project administration, manuscript revisions, and its final approval; SPZ and HYW, formal analysis, manuscript revision, and final approval; CYF, HYL and SY, conception, manuscript revision, and final approval; and NS, conception, design, funding acquisition, project administration, manuscript revision, and final approval. All the authors have read and approved the final manuscript.

## Acknowledgments

The authors would like to thank the older adults who participated in the study. The authors also thank the source of financial grants: Zhejiang Philosophy and Social Science Planning Project and Zhejiang Province Universities Major Humanities and Social Sciences Tackling Plan Project.

